# The effect of artificial oocyte activation on blastocysts rate in patients with low blastocyst rates: A retrospective cohort study

**DOI:** 10.1101/2024.06.28.24309669

**Authors:** Feras Sendy, Robert Hemmings, Isaac-Jacques Kadoch, Wael Jamal, Simon Phillips

## Abstract

**Introduction:** Physiological oocyte activation requires a synergy between the oocyte and sperm to release calcium (Ca2+) through oscillations. The absence of such synergy between the oocyte and sperm leads to a negative impact on oocyte activation. Studies have shown that Artificial oocyte activation (AOA) is helpful in cases with failed or low fertilization rates. Studies present mixed opinions about increasing blastocyst rate.

**Methods:** A retrospective cohort single-center study was performed between January 2018 and October 2023, including 54 couples with suboptimal blastocyst development. The study compared intracytoplasmic sperm injection (ICSI) AOA cycles with previous conventional ICSI cycles and conventional ICSI without AOA cycles with previous conventional ICSI cycles in couples with failed or low blastocyst rates (< 30%) in the original ICSI cycle.

**Results:** We compared 22 AOA cycles to previous conventional ICSI cycles in the same patients and 32 conventional ICSI cycles without AOA to previous conventional ICSI cycles in the same patients. After AOA, the blastocyst rate was not significantly higher than the control group (48% vs 29% p=0.19). Conversely, the blastocyst rate was significantly higher in the conventional ICSI without AOA cycles than in the control group (48% vs 24% p=0.04). The fertilization rate was not statistically significant between the first and second cycles in both groups.

**Conclusion:** The literature still lacks strong evidence for AOA overcoming impaired embryonic development. Therefore, AOA remains reserved for couples with a failed or low fertilization history to improve fertilization results. Optimal laboratory conditions and ovarian stimulation modifications without AOA may improve blastocyst rates.

## Introduction

Infertility affects males and females for various reasons (1). Currently, 7% of all births have been achieved by assisted reproductive technologies (ART) (2). Intracytoplasmic sperm injection (ICSI) is a technique used in the case of male infertility causes and following failure of in vitro fertilization (IVF), giving successful fertilization in 70% of the cases (2). However, certain infertile patients cannot achieve adequate fertilization due to the absence of synergy between the sperm and oocyte to release calcium (Ca2+) in the form of oscillations (2, 3). Globozoospermia is one of the causes that leads to an absence of such synergy due to the abnormal formation or absence of the sperm acrosome (4). Moreover, globozoospermia impacts oocyte activation due to PLCζ abnormal expression, localization, and protein structure (5, 6).

Artificial oocyte activation (AOA) can be carried out by mechanical, chemical, or electrical means (7). Mechanical manipulation of the injecting pipette during ICSI is the least invasive and least effective method (8). Electrical methodology, by direct voltage, allows extracellular Ca2+ entry, however it has a high oocyte degeneration rate (9). Thus, using chemical components such as calcimycin or ionomycin for AOA became the most common method (7). Calcimycin or ionomycin has been linked with success in improving the fertilization rate (10, 11). The European Society of Human and Reproduction (ESHRE) recommends AOA in case of fertilization failure, low fertilization (< 30%), or globozoospermia (12).

AOA has been proposed as a possible treatment for embryo development after ICSI (13-15). Nevertheless, studies have presented mixed opinions concerning increases in the blastocyst and live birth rates (13-15). Furthermore, each study had different inclusion criteria, study design, and Ca2+ ionophore use during AOA.

The present study investigated whether AOA using ionomycin can improve blastocyst rate in patients with poor or no blastocyst development on their previous IVF-ICSI attempt. Secondary objectives were the percentage of patients without usable embryos, fertilization, and pregnancy rates.

## Material and Methods

### Study design, setting, and participants

A retrospective cohort single-center study was performed between January 2018 and October 2023, including 54 couples with a history of poor blastocyst development (< 30%) in a previous ovarian stimulation cycle. Group 1 consists of 22 patients who underwent an ICSI-AOA cycle following a failed conventional ICSI cycle. Group 2, 32 patients, underwent a second conventional ICSI cycle without AOA following a failed conventional ICSI cycle. Group 2 represents a control group since failed cycles are presented to a multidisciplinary medical council at our clinic to assess options to improve outcomes on a second cycle. Modifications to the ovarian stimulation protocol including changes of gonadotropins, use of adjuvants, or ovulation trigger medications are often prescribed. Comparing any improvements seen in the AOA group to those seen in this control group, reduces the impact of other modifications to treatment carried out in addition to the AOA. Of the 54 couples included, 22 underwent an ICSI-AOA cycle consisting of calcium ionophore (GM508-CultActive) exposure, and 32 couples underwent conventional ICSI without an AOA cycle. The flow chart illustrates the included and excluded cases (Figure 1). The time between the first and second stimulation cycles ranged between 15 days – 4 years. The mean maternal age ranged between 35 -36 years old. Couples were diagnosed with male factor infertility, low ovarian reserve, unexplained female infertility, polycystic ovarian syndrome (PCOS), and genetic disease.

**Figure 1:**
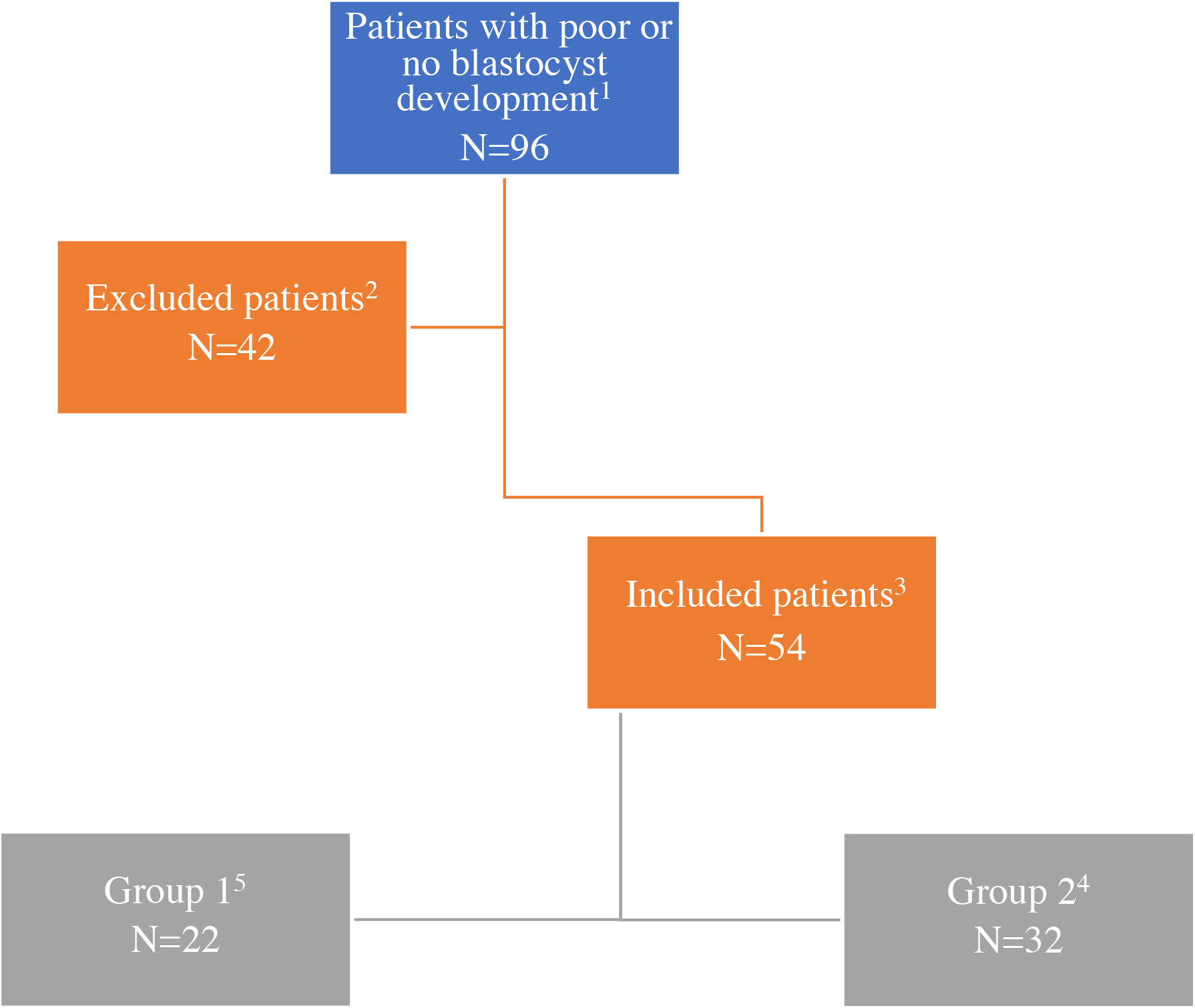
Flowchart explaining the number of included and excluded couples. 1: Patients with stimulation cycles that resulted in poor or no blastocysts (< 30%). 2: Patients with absent comparable cycles (without a previous conventional ICSI cycle, second conventional ICSI cycle, or second ICSI -AOA). 3: Patients with comparable cycles. 4: Patients with a first conventional ICSI cycle and a second ICSI-AOA cycle. 5: Patients with a first conventional ICSI cycle and a second conventional ICSI cycle without AOA.

### ICSI and ICSI-AOA procedures

Female partners were mainly stimulated using an antagonist protocol. Some cases were stimulated using a short agonist protocol. Ovulation was triggered by administering human chorionic gonadotropin (hCG), gonadotropin-releasing hormone agonist (GnRH agonist), or both. At 36-h post ovulation trigger administration, oocytes were aspirated from the follicles in the ovaries by transvaginal ultrasound-guided puncture in Global Total™ medium (Cooper Origio, USA). Oocyte denudation was carried out using Cumulase™ (Coper-Origio, USA), and oocytes were assessed for maturity before ICSI. Sperm selection was performed using density gradient centrifugation or Zymot™ and fresh or frozen-warmed sperm was used. Only motile spermatozoa were immobilized before injection into mature oocytes.

For AOA, oocytes were exposed to Gynemed GM508 Cultactive (Fertitech Canada) for 15 minutes post-ICSI, washed through Global Total™ with HEPES, and cultured in Global Total™.

For Both ICSI and ICSI-AOA cycles, embryo culture was performed in Global Total™, and embryo transfer was performed fresh on day five or in a frozen embryo transfer cycle using a blastocyst.

### Statistics

Statistical analysis was performed using IBM SPSS version 26. Following assessment of the normal distribution for continuous independent variables, means were expressed with standard deviations and compared by T-test or Anova or linear regression.

Independent binary variables including Maturation(MII/Oocytes), fertilization (2PN/MII), blastocyst development (blastocyst/2PN), and pregnancy rate (HCG+/transfer) were compared between the first and second cycles in the two groups by Chi-Square test or logistic regression. An alpha error of 0.05 was considered significant.

P-values < 0.05 were considered as statistically significant.

## Results

For Group 1, the blastocyst rate was not significantly higher with AOA than conventional ICSI without AOA (48% versus 29%, p= 0.19). However, the clinical pregnancy rate was significantly higher with AOA than conventional ICSI without AOA (45% versus 13%, p=0.01). Fertilization and maturation rates were not statistically significant. The percentage of patients without usable embryos was not significantly lower with AOA compared to the conventional ICSI cycle without AOA. All the results for this group are demonstrated in Table 1.

**Table 1:** Results of Group 1 patients (second stimulation cycle with ICSI-AOA following a previous conventional ICSI cycle without AOA.) *: Statistically significant with a P value of < 0.05

|  | **Cycle1 : Conventional**<br>**ICSI**<br>**(N=22)** | **Cycle 2 : ICSI -AOA**<br>**(N=22)** | **P value** |
| --- | --- | --- | --- |
| **AMH** | 3.4±3.2 | 3.3±3.2 | 0.91 |
| **Maturation rate** | 11±7.3 | 12±10 | 0.70 |
| **Fertilization rate** | 53% | 60% | 0.34 |
| **2PN/Patient** | 6.2±5.9 | 7.3±8.1 | 0.60 |
| **Blastocyst number** | 1.8±2.5 | 3.5±3.0 | 0.04* |
| **Blastocyst rate** | 29% | 48% | 0.19 |
| **% of patients without usable**<br>**embryos** | 54% | 27% | 0.06 |
| **Pregnancy rate** | 3/22 (13%) | 10/22 (45%) | 0.01* |

In Group 2, the blastocyst rate was significantly higher with conventional ICSI without AOA than previous conventional ICSI (48% versus 24%, p=0.04). The clinical pregnancy rate was significantly higher with conventional ICSI without AOA than previous conventional ICSI (53% versus 28%, p=0.04). However, the maturation and fertilization rates were not statistically significant. The percentage of patients without usable embryos was not significantly lower with the second conventional ICSI cycle than with their previous conventional ICSI cycle. The outcomes for this group are demonstrated in Table 2. Comparative analysis of pregnancy, blastocyst, and fertilization between group 1 and group 2 second cycles are summarized in Table 3.

**Table 2:** Results of Group 2 (second stimulation cycle with conventional ICSI without AOA following a previous conventional ICSI cycle.) *: Statistically significant with a P value of < 0.05

|  | **Cycle 1: Conventional**<br>**ICSI**<br>**(N=32)** | **Cycle 2: Conventional**<br>**ICSI**<br>**(N=32)** | **P value** |
| --- | --- | --- | --- |
| **AMH** | 3.1±2.6 | 3.0±2.5 | 0.87 |
| **Maturation rate** | 7±4.6 | 8±5 | 0.40 |
| **Fertilization rate** | 72% | 68% | 0.72 |
| **2PN/Patient** | 4.9±3.6 | 5.9±3.7 | 0.27 |
| **Blastocyst number** | 1.2±1.8 | 2.8±2.3 | 0.001* |
| **Blastocyst rate** | 24% | 48% | 0.04* |
| **% of patients without usable**<br>**embryos** | 43% | 12% | 0.005* |
| **Pregnancy rate** | 8/28 (28%) | 15/28 (53%) | 0.04* |

**Table 3:** Results from the second cycles of Group 2 and Group 1. *: Statistically significant with a P value of < 0.05

|  | **Group 2**<br>**(N=32)** | **Groupe 1**<br>**(N=22)** | **P value** |
| --- | --- | --- | --- |
| **Percentage of fertilization** | 68% | 60% | 0.54 |
| **Percentage of Blastocyst** | 48% | 48% | 1.0 |
| **Percentage of Pregnancy** | 53% | 45% | 0.54 |

## Discussion

In this study, we showed that AOA treatment neither improved the blastocyst rate nor the pregnancy rate in our patient cohort with previous poor blastocyst rates (< 30% blastocyst formation). The findings were not influenced by fertilization, as both cycles had comparable rates. Although, the cycle-to-cycle duration effect was not measured we believe that the duration between cycles does not seem to impact results given that no significant changes occurred in the laboratory culture system during the period under evaluation. Furthermore, any generalized improvements would have primarily affected the second cycle of the AOA group, adding more weight to the findings that AOA did not improve the blastocyst development rate.

There is evidence that ICSI-AOA improves fertilization rates leading to blastocysts in patients with poor fertilization (16-18). Oocyte activation is impacted by the sperm or oocyte due to the absence of synergy to produce Ca2+. As a result, AOA during fertilization helps conquer this issue. Other studies did not find an improved effect (19, 20). AOA has been proposed for embryo development after ICSI, yet the results remain mixed (13-15). For example, Ebner et al. (2015) used GM508 Cult-Active to treat 57 patients with reduced blastocyst rate (≤15%) and found a significant increase in blastocyst and pregnancy rates after AOA. Similarly, our study found a significant increase in pregnancy rate in group 1 (Table 1). However, the blastocyst rate was not statistically significant.

Similarly, results from (14, 15) did not find AOA’s advantage in improving blastocyst rate. Yin et al. (2022) used ionomycin to treat 140 patients with reduced day three embryo rate (≤30%) and found no difference in day three embryo, blastocyst, and pregnancy rates. The inclusion criteria of poor blastocyst rate (≤30%) in our study was different from Yin et al. (2022) and Ebner et al. (2015) who used cutoffs of poor cleavage stage (≤30%) and poor blastocyst stage (≤15%).

Mateizel et al. (2022) used calcimycin to treat 15 patients with poor blastocyst rate and found no difference in blastocyst rate. Both studies (14, 15) used different Ca2+ ionophores (calcimycin versus ionomycin), and embryo stages (blastocyst versus cleavage). Nonetheless, they used sibling oocytes as a control group and found similar outcomes. Moreover, Ebner et al. (2015) compared ICSI-AOA to the previous ICSI cycle with poor blastocyst rate, a similar criterion in our study.

The lack of Ca2+ oscillations during fertilization is one of the reasons for fertilization failure. A study found that AOA may develop higher activation using ionomycin rather than calcimycin (21). Some patients with failed ICSI-AOA by calcimycin may be rescued by ionomycin (22). Thus, much higher stimuli is required for some patients than others (23). We could not examine the effect of using different ionophores or sibling oocytes. However, our study compared group 1 (Table 1), group 2 (Table 2), and both groups (Table 3) but did not find a significant impact on fertilization rate.

Our study did not evaluate the miscarriage or live birth rate. However, the literature provides evidence of AOA’s effectiveness and safety when used in adequate conditions (fertilization failure or low fertilization). A meta-analysis of 14 studies found that AOA increased live birth, and pregnancy rates with no significant effect on miscarriage rate(18). Furthermore, globozoospermia is an indication of AOA by ionomycin than calcimycin (30% versus 11.8% respectively)(11). Chromosomal abnormalities, gene expression, or morpho kinetics due to Ca2+ ionophores are unlikely due to the inability of Ca2+ to enter the oocyte (24, 25). Moreover, various studies reported the absence of any link between AOA and birth defects (26-28). Likewise, cognition, language, and motor skills were normal in children (29).

The stimulation protocol and dosage choice depend on the patient’s age, weight, ovarian reserve, and response to previous stimulation. The use of antagonist stimulation protocol resulted in a comparable ongoing pregnancy rate to agonist stimulation protocol for poor responders in a meta-analysis involving 50 studies (30). A randomized controlled trial looked at the impact of constant versus increased stimulation dosage under antagonist protocol (31). The number of oocytes retrieved, fertilization, and pregnancy rates were not significantly different. Furthermore, a systematic review of thirty studies compared stimulation with recombinant follicular stimulating hormone (rFSH) and rFSH with recombinant luteinizing hormone (rLH) (32). They found that hypo responders and advanced age (36-39 years old) might have better oocyte retrieval and implantation rates in stimulation using rFSH and rLH. However, a randomized controlled trial in 240 patients over thirty-five years old did not find a significant live birth rate in rFSH and rLH compared to rFSH alone (33). Moreover, a meta-analysis of 22 studies found comparable clinical pregnancy rates yet lower consumption of FSH in poor responders with clomiphene citrate or aromatase inhibitors during stimulation in comparison to ovarian stimulation without the addition of clomiphene citrate or aromatase inhibitors (34). Ovulation induction using an agonist or HCG showed a comparable number of oocytes retrieved and live birth rate in a randomized controlled trial for 257 women undergoing oocyte donation (35). Using double triggers (agonist and HCG) for ovulation induction has been advocated for increasing oocyte maturation (36). Most of our patients had an antagonist stimulation protocol with or without aromatase inhibitors, and some of them had ovulation trigger using the agonist alone or in combination with HCG depending on the risk of ovarian hyperstimulation syndrome. Others had short agonist stimulation as a change of previous stimulation, poor response, or other reasons. The results of the second stimulation cycle in conventional ICSI and ICSI-AOA have improved compared to the first. The improvement could be attributed to the change of stimulation protocol, the inadequate dosage of stimulation in the first cycle, or the absence of a double trigger in some cases.

Our data show that AOA is not indicated for patients with a history of poor blastocyst rate after ICSI (< 30%) as it probably results from other factors not related to Ca2+. Despite our low sample size, we have shown that AOA does not improve blastocyst rate since Group 2 patients improved on their second conventional ICSI cycle without this intervention being applied. This technique should be reserved for patients with failed or low fertilization (12). Optimizing laboratory conditions and ovarian stimulation protocols may improve blastocyst rates for such patients.

### Limitations

Our strict inclusion criteria, requiring at least one previous ICSI cycle with a failed or low blastocyst development rate, permitted an ideal comparison of the same patient groups. However, this criterion limited the sample size of the study.

## Conclusion

The literature lacks strong evidence for AOA overcoming impaired embryonic development. This study suggests no additional benefit to using AOA for patients with previous poor blastocyst rates. Therefore, AOA should be reserved for couples with failed or low fertilization history to improve fertilization results. Optimal laboratory conditions with/without AOA may lead to improved blastocyst rates.

### What is known about this topic

- Certain infertile patients are unable to achieve adequate fertilization due to the absence of synergy between the sperm and oocyte to release calcium in the form of oscillations.
- AOA can be carried out by mechanical, chemical, or electrical means.
- AOA has been proposed as a possible treatment for embryo development after ICSI. Nevertheless, studies have presented mixed opinions concerning increases in the blastocyst and live birth rates.

### What this study adds

- AOA is reserved for couples with a history of failed or low fertilization (< 30%) to improve fertilization results.
- Adequate laboratory conditions and ovarian stimulation may improve blastocyst rates.

## Data Availability

All data produced in the present work are contained in the manuscript

## Ethical approval

Ethical approval for the study was obtained from the relevant local ethics committees.

## Author’s contributions

Feras Sendy served as the lead author, contributing to all stages of the research, from study design to paper writing. Robert Hemmings contributed to statistical analysis, interpretation, and review the paper. Wael Jamal and Isaac-Jacques Kadoch contributed to interpretation, review, and validation of the paper. Simon Phillips contributed to interpretation, review, and validation of the paper from study design to paper writing.

